# Uncoupling interferons and the interferon signature explain clinical and transcriptional subsets in SLE

**DOI:** 10.1101/2023.08.28.23294734

**Authors:** Eduardo Gómez-Bañuelos, Daniel W. Goldman, Victoria Andrade, Erika Darrah, Michelle Petri, Felipe Andrade

**Author notes:** Address for correspondence and reprints: Eduardo Gómez-Bañuelos, 5200 Eastern Avenue, Mason F. Lord Building Center Tower, Room 676 Baltimore, Maryland, 21224, USA, Felipe Andrade, 5200 Eastern Avenue, Mason F. Lord Building Center Tower, Room 663 Baltimore, Maryland, 21224, USA.

## Abstract

Interferons (IFN) are thought to be key players in systemic lupus erythematosus (SLE). The unique and interactive roles of the different IFN families in SLE pathogenesis, however, remain poorly understood. Using reporter cells engineered to precisely quantify IFN-I, IFN-II and IFN-III activity levels in serum/plasma, we found that while IFNs play essential role in SLE pathogenesis and disease activity, they are only significant in specific subsets of patients. Interestingly, whereas IFN-I is the main IFN that governs disease activity in SLE, clinical subsets are defined by the co-elevation of IFN-II and IFN-III. Thus, increased IFN-I alone was only associated with cutaneous lupus. In contrast, systemic features, such as nephritis, were linked to co-elevation of IFN-I plus IFN-II and IFN-III, implying a synergistic effect of IFNs in severe SLE. Intriguingly, while increased IFN-I levels were strongly associated with IFN-induced gene expression (93.5%), in up to 64% of cases, the IFN signature was not associated with IFN-I. Importantly, neither IFN-II nor IFN-III explained IFN-induced gene expression in patients with normal IFN-I levels, and not every feature in SLE was associated with elevated IFNs, suggesting IFN-independent subsets in SLE. Together, the data suggest that, unlike the IFN signature, direct quantification of bioactive IFNs can identify pathogenic and clinically relevant SLE subsets amenable for precise anti-IFN therapies. Since IFN-I is only elevated in a subset of SLE patients expressing the IFN signature, this study explains the heterogeneous response in clinical trials targeting IFN-I, where patients were selected based on IFN-induced gene expression rather than IFN-I levels.

## Introduction

Systemic lupus erythematosus (SLE) is a complex autoimmune disease affecting multiple organs, which is characterized by marked clinical heterogeneity, a fluctuating course with relapses and remissions, and high titer antibodies to diverse autoantigens.^1^ While the etiology of SLE remains unknown, bulk and single cell transcriptomic profiling of peripheral blood and target tissues from patients with SLE have identified unique transcriptional signatures associated with dysregulation of immune-related pathways, including a prominent transcriptional profile linked to increased signaling by interferons (IFNs).^2–10^ The IFN family comprises type I IFNs (IFN-I, including 12 IFNα subtypes plus IFNβ, IFNɛ, IFNκ and IFNω), type II IFN (IFN-II, IFN-γ) and type III IFNs (IFN-III, IFN-λ1-4).^11^ Multiple studies have shown that circulating levels of members of all three families are elevated in SLE, and overall, increased levels of IFNs have been associated with higher disease activity and some IFN subtypes with distinct clinical features.^12–16^ The individual and interacting contributions of the different IFN families in SLE pathogenesis, however, remain poorly understood.

A significant problem in the study of IFNs in SLE has been defining the best method for quantification. Except for IFN-II, which only includes IFN-γ, IFN-I and IFN-III comprise multiple subtypes, making it difficult to quantify them individually or in bulk with a single assay. While new technologies have been used to overcome this limitation and measure the majority of IFN-α’s in SLE serum/plasma using a single assay (e.g., single-molecule array digital ELISA technology),^16^ other IFN-I members that may have functional relevance in SLE were excluded (e.g., IFNβ, IFNɛ, IFNκ and IFNω). More importantly, not all IFN-α subtypes have the same effect to activate cells. According to their potency, IFN-α’s can be classified as having low (IFN-α1), intermediate (IFN-α2a, –4a, –4b, –5, –16, –21), and high (IFN-α2b, – 6, –7, –8, –10, –14) activity.^17^ Since IFN-α subset levels are likely to vary among SLE patients and over time, it is expected that similar circulating amounts of total IFN-α will have variable activities according to the proportion of IFNs with low, intermediate and high activity. This notion explains why bulk IFN-α levels and activity do not correlate in a significant number of healthy individuals and SLE patients.^16,18^ Therefore, quantifying total levels of IFN-α may not necessarily reflect the systemic activity of IFN-I in patients with SLE.

Regarding other IFNs, the relationship between IFN-III levels and SLE is likely among the most inconsistent, including divergences in the association with disease activity, clinical features, and the subset of elevated IFN-III (i.e., λ1, λ2 or λ3).^15,19–23^ These discrepancies are most likely due to differences in the methods used to quantify IFN-III and importantly, the subtype of IFN selected as “exemplary” to reflect levels of all four IFN-III.

Because of the difficulties in directly assessing circulating levels of IFNs, alternative approaches have been used to study the effect of IFNs in SLE. One method consists in correlating IFN activity with SLE by estimating differentially expressed IFN-stimulated genes (ISGs) in patient-derived cells, which is known as the IFN signature.^4,5,24,25^ This transcriptional profile, however, has a significant overlap among members of all IFN families,^26–28^ limiting the IFN signature as a method to define the independent contribution of the different IFN families to SLE. Other commonly used methods are functional activity assays, where reporter cells are exposed to serum/plasma and the IFN activity is determined by the transcriptional expression of ISGs or by a reduction in the cytopathic effect of viruses.^12,16,29^ Since members of the same IFN family can only signal through a single receptor,^30^ this method is convenient because it reports on the coordinated activity of members of a single IFN family. Therefore, functional assays are more precise than quantifying bulk or individual amounts of IFN subtypes in capturing the effector function of circulating IFNs at the cellular level in SLE. Functional IFN assays, however, have significant limitations. They are laborious, have only been applied for the study of IFN-I,^12,29,31^ and their specificity may require blocking IFN-III because reporter cells are not IFN-I specific.^32,33^ Here, to investigate the relationship between IFN families – individually and collectively – with clinical and transcriptional profiles in SLE, we used a novel and convenient cell-based reporter assay to quantify the specific activity of IFN-I, IFN-II, and IFN-III in combination with clinical and whole blood transcriptional data from a large prospective cohort of patients with SLE.

## Results

### Activity levels of IFN-I, IFN-II and IFN-III are elevated in SLE

Activity levels of human IFN types were quantified using commercial reporter cells engineered to express specific receptors and transcription factors to precisely detect bioactive human IFN-I, IFN-II, or IFN-III (InvivoGen). Similar to experimental data provided by the manufacturer, we validated that each cell line is specific for detecting its corresponding type of IFN (**figure 1A-C**). Moreover, using matched SLE serum and plasma, IFN activity levels showed a significant correlation (IFN-I, r^2^ = 0.866, P < 0.0001; IFN-II, r^2^ = 0.777, P < 0.0001; IFN-III, r^2^ = 0.745, P < 0.000.1; **Supplemental figure 1**), indicating that blood processing has minimal effect on IFN concentrations, which is consistent with previous reports.^16^

**Figure 1.**
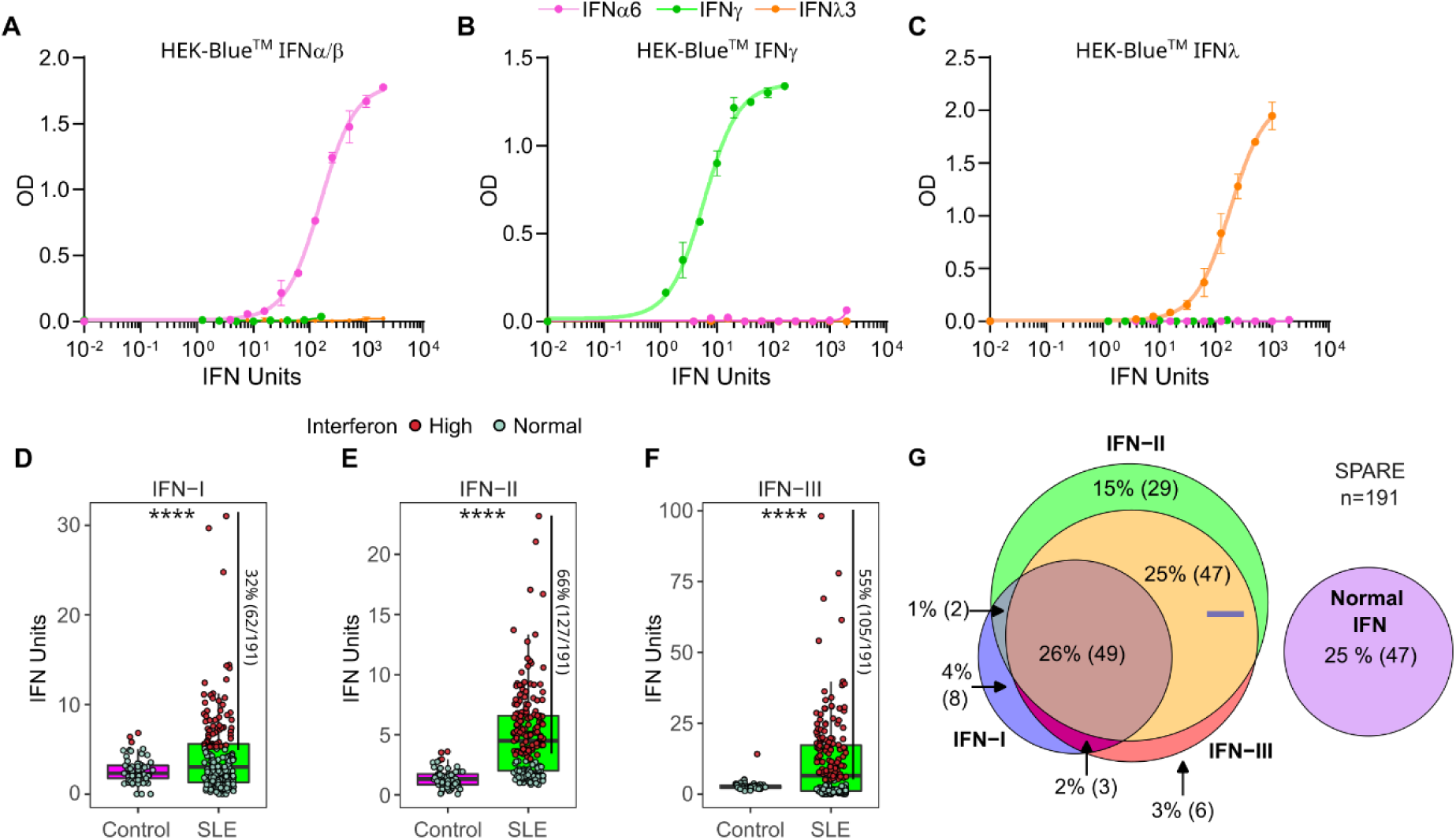
Activity levels of IFN-I, IFN-II, and IFN-III in patients from the SPARE cohort. A-C. HEK-Blue^TM^ IFN-α/β (**A**), HEK-Blue^TM^ IFN-γ (**B**) and HEK-Blue^TM^ IFN-λ (**C**) cells were incubated with increasing amounts of IFN-α6, IFN-γ and IFN-λ3. The cells only detected their specific IFN with a range of 1.0 to 2000 IU for IFN-I and IFN-III, and from 1 to 160 IU for IFN-II. **D-F.** Activity levels of IFN-I, IFN-II and IFN-III in SLE and healthy controls (HC). Samples with elevated levels of IFN-I, IFN-II or IFN-III are colored in red. Cut-off was determined by ROC curve analysis with a 95% specificity vs. HC. Elevated levels of IFN were set at 5.17 U, 2.9 U and 5.0 U for IFN-I, IFN-II, and IFN-III, respectively. In patients with serial samples, only the first sample was included in the analysis. p values were obtained using Student’s t test. ****p < 0.0001. **G.** Venn diagram showing the intersection between elevated IFN types in patients with SLE.

Circulating activity levels of IFN types were then determined in 56 healthy controls and 191 SLE patients from the “Study of biological Pathways, Disease Activity and Response markers in patients with SLE” (SPARE).^34–36^ Demographic, clinical, and laboratory features of the SLE cohort are summarized in **Supplemental table 1**. IFN activity was detected both in healthy control and SLE samples (**figure 1D-F**). Compared to healthy controls, however, patients with SLE showed significantly higher activity of IFN-I, IFN-II, and IFN-III [mean units (SD), 2.6 (1.5) vs. 4.3 (4.5), 1.4 (0.8) vs. 5.0 (3.5), and 2.9 (1.7) vs. 11.3 (14.4), respectively, p < 0.0001 in all cases] (**figure 1D-F**). Using a ROC curve to determine cut-off points for each IFN, elevation in IFN-II was the most prominent in SPARE (66%, 127/191), followed by IFN-III (55%, 105/191), and IFN-I (32%, 62/191) (**figure 1D-F**). Moreover, 22% (43/191) of SLE patients showed elevation of only one type of IFN, 53% (101/191) had more than one type of IFN elevated, and 25% (47/191) had IFN activities within the range of healthy controls (termed normal IFN) (**figure 1G**).

### Activity levels of IFN types have different patterns according to disease activity and disease duration

To analyze the stability of the IFN-types over time in SLE, we assessed the activity of the three IFN families in 226 longitudinal samples from 78 patients, and calculated the intraclass-correlation coefficient (ICC) to measure ‘within’ subject variability or repeatability using a linear mixed-effects model^37^ (**figure 2A**). During follow-up, IFN-I exhibited the greatest repeatability among the three IFN types, followed by IFN-II and IFN-III [ICC (95% CI), 0.529 (0.381, 0.648), 0.369 (0.224, 0.509) and 0.235 (0.085, 0.348)]. Overall, all IFN types showed lower ICC compared to body weight, which was the most stable parameter during follow-up [ICC (95% CI), 0.975 (0.962, 0.982) p < 0.0001], but comparable to SLEDAI [ICC (95% CI), 0.319 (0.169, 0.464)] (**figure 2A**), suggesting that similar to disease activity, IFN levels are highly variable during the course of SLE.

**Figure 2.**
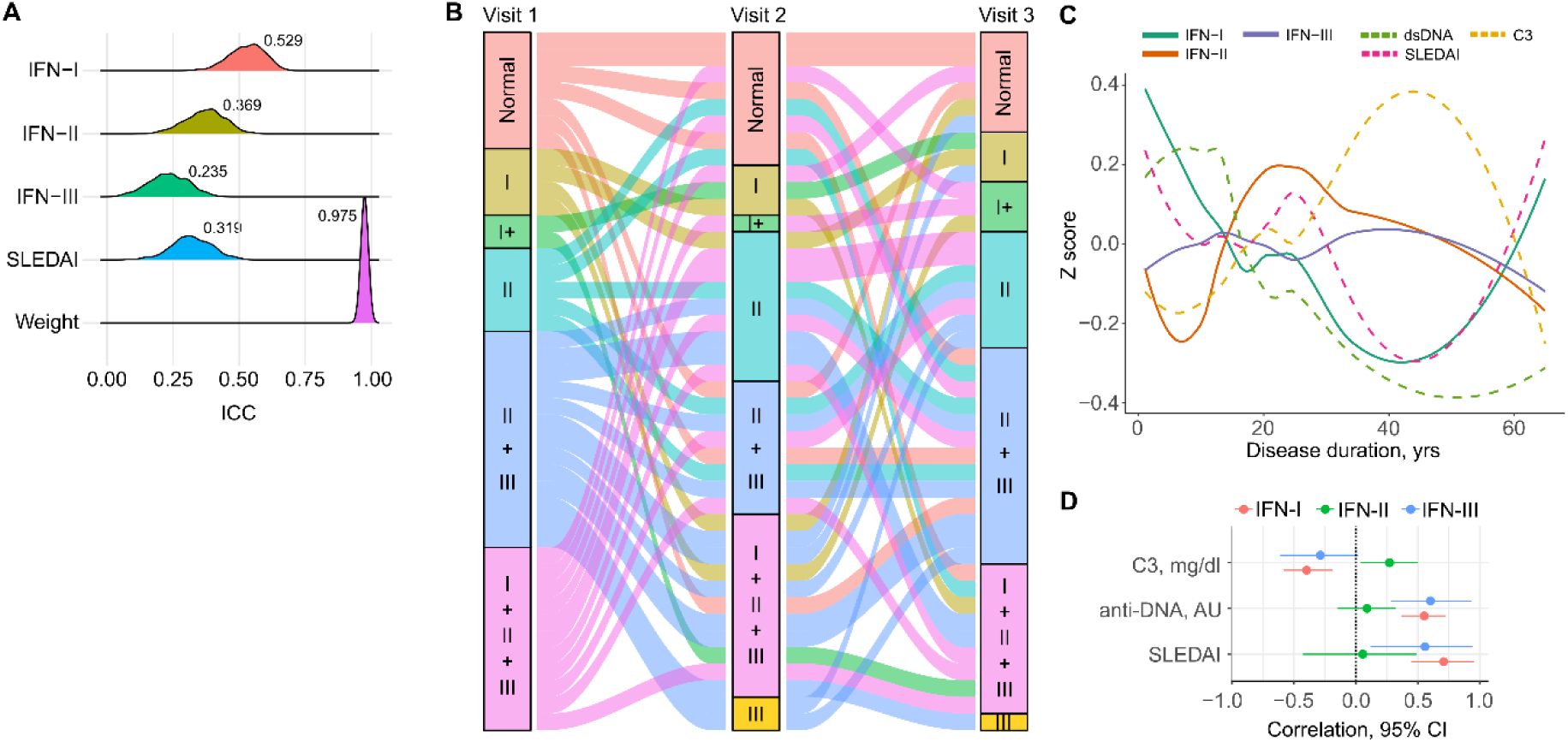
**Longitudinal patterns of IFN type and their association with markers of disease activity in SLE. A**. Intra class-correlation coefficient (ICC) of IFN activity levels in SLE patients with more than 1 longitudinal sample (n=77). **B**. Sankey diagram showing the flow of IFN levels in SLE patients with three longitudinal samples (n=42). Normal = Normal IFN; I = High IFN-I; I+ = High IFN-I plus high IFN-II or IFN-III; II = High IFN-II; III = High IFN-III; II+III = High IFN-II + IFN-III; I+II+III = elevation of the three IFN types. Node height represents the proportion of SLE patients with the indicated elevation of IFNs at the time of visit. Visit 1: Normal, 17% (n=7); I, 10% (n=4); I+, 5% (n=2); II, 12% (n=5); II+III, 31% (n=13); I+II+III, 26% (n=11). Visit 2: Normal, 18% (n=8); I, 7% (n=3); I+, 2% (n=1); II, 21% (n=9); II+III, 19% (n=8); I+II+III, 26% (n=11), III, 5% (n=2). Visit 3: Normal, 14% (n=6); I, 7% (n=3); I+, 3% (n=7); II, 7% (n=17); II+III, 31% (n=13); I+II+III, 21% (n=9); III, 2% (n=1). Each flow or link between the nodes represents a single patient. **C**. Trajectory of the different IFN types along with SLEDAI, C3, C4 and anti-dsDNA in patients with SLE. To represent grouped trajectories, z-scores of IFN type and disease activity markers (SLEDAI, C3, and dsDNA) were projected over disease duration using loess-fitted curves. **D**. Correlation between disease activity markers and IFN type obtained by a Bayesian mixed-effects model in patients with SLE adjusted by disease duration.

To better depict the stability of IFNs over time, we selected a subset of SLE patients (n = 42) who had IFN measurements in three consecutive visits (**figure 2B**). Because the majority of the patients had elevated levels of more than one IFN type, they were divided into subsets based on the single or combined activity of the three IFN types. Ninety percent (38/42) of SLE patients changed their IFN subset through the three visits. Among these, 72% of patients with normal IFN levels at baseline developed a single or combined elevated IFN activity in the subsequent visits (**figure 2B**), confirming that IFN subsets are highly dynamic over the course of the disease.

To determine whether the variability in IFN activity was associated with disease activity, we fitted a linear mixed-effects model using data from all SLE patients. IFN-I and IFN-III were significantly associated with SLEDAI [exp(β) (95% CI), 1.06 (1.04,1.08) and 1.06 (1.02,1.1), respectively], IFN-II with longer disease duration, and IFN-I was negatively associated with disease duration [exp(β) (95% CI), 1.01 (1.0,1.01) and 0.99 (0.98,0.99), respectively] (**Supplemental table 2-4**). To better understand the relationship between IFN activity levels and disease activity over time, we projected the IFN levels and disease activity parameters over disease duration (**figure 2C**). Notably, in contrast to IFN-II and IFN-III, IFN-I levels paralleled changes in SLEDAI, anti-dsDNA and C3 (**figure 2C**). Using a bayesian mixed-effects model to examine the correlation of individual IFN types with disease activity measures (**figure 2D**), we confirmed that changes in IFN-I were linked with changes in SLEDAI, anti-dsDNA and C3. In contrast, changes in IFN-II were associated with increased levels of C3, and IFN-III was mildly correlated with changes in SLEDAI and anti-dsDNA.

### IFN types define distinct clinical subsets in SLE

To determine clinical endotypes associated with IFNs, we analyzed the relationship between SLEDAI specific items and IFN activity levels (**Supplemental Table 5**). Low complement, anti-DNA antibodies, rash, and proteinuria were predictive of higher IFN-I activity, while mucosal ulcers were associated with lower IFN-I activity (**figure 3A**). Arthritis and recent bacterial infections were associated with increased IFN-II (**figure 3B**), and oral candida, low complement, and renal involvement were predictive of increased IFN-III activity (**figure 3C**).

**Figure 3.**
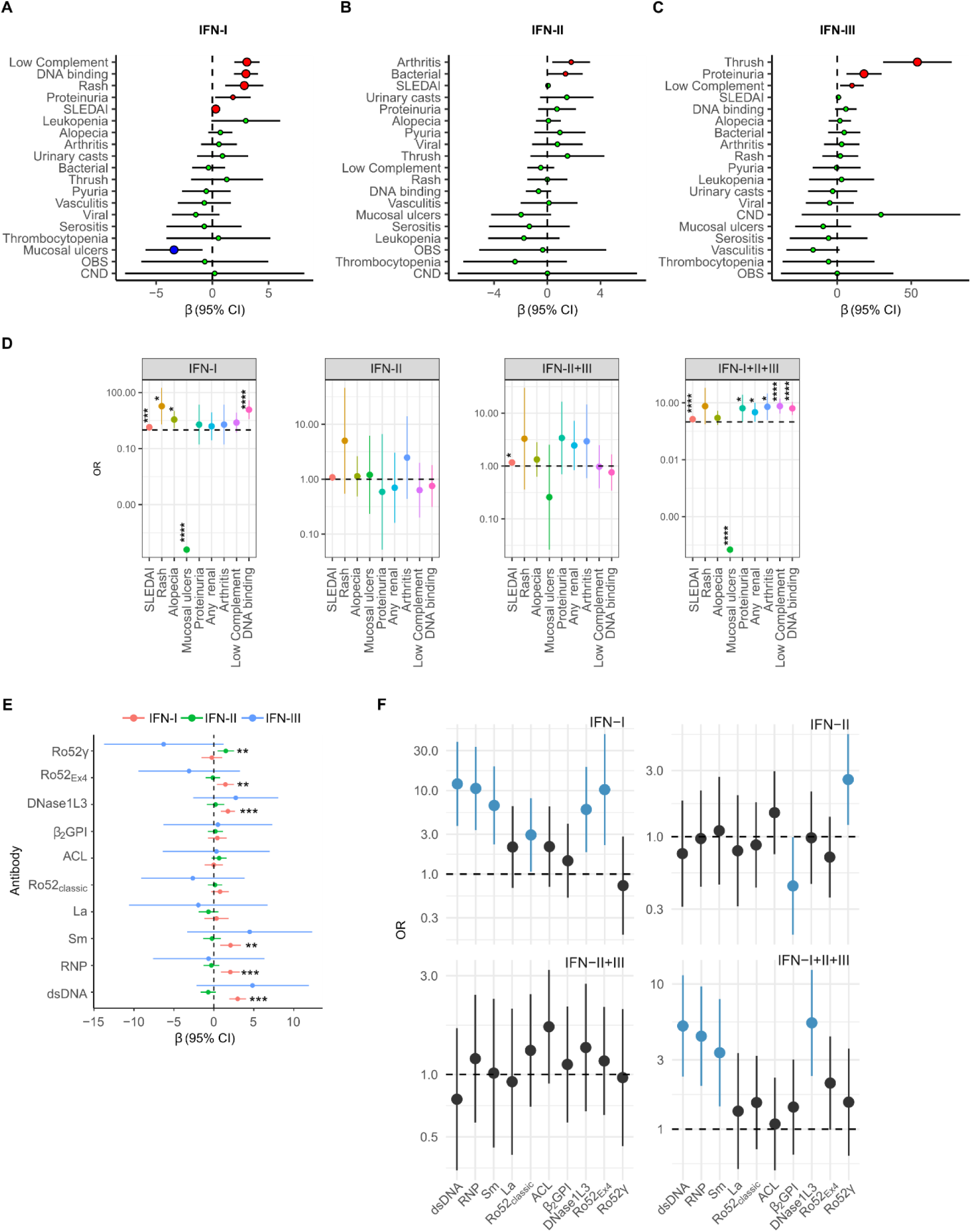
SLE disease subsets are predictive of increased activity levels of IFN types. A-C. Effect of SLE activity features at time of visit on levels of IFN-I (**A**), IFN-II (**B**), and IFN-III (**C**). **D**. Predictive value of disease activity items for single or combined IFN types. **E.** Effect of autoantibodies in SLE over the IFN types. **F.** Predictive value of autoantibodies over individual or combined IFN type in SLE. In **A**-**C** and **E**, associations were evaluated using a mixed-effects linear regression model adjusted by disease duration, considering IFN activity as dependent variable. In **D** and **F**, associations were evaluated using a mixed-effects multinomial logistic regression adjusted by disease duration, considering the IFN groups as dependent variables, SLEDAI items or autoantibodies as predictors, and the group with normal IFN as reference. *p<0.05, **p< 0.01, ***p<0.001, ****p<0.0001.

Given that most patients exhibit elevated levels of more than one IFN type (**figure 1G**), a multinomial logistic regression model was used to evaluate the predictive value of disease activity score items across individual and combined IFN groups (**Supplemental table 6**). SLEDAI was predictive of the single elevation of IFN-I, as well as combined IFN activity (i.e., IFN-II + IFN-III and IFN-I + IFN-II + IFN-III) (**figure 3D**). Single elevation of IFN-I was associated with skin manifestations (rash and alopecia) and anti-DNA binding, while IFN-II elevation alone was not associated with any disease activity item (**figure 3D**). Interestingly, features of systemic disease – such as arthritis, nephritis and low complement – were only significantly associated with co-elevation of the three IFN types (**figure 3D**), suggesting an additive effect of IFNs in severe SLE. Other IFN groups (i.e., IFN-I + IFN-II, IFN-I + III, and IFN-III alone) were not included in this analysis due to the small number of patients in each group.

Except for antibodies to Ro52, antibodies to dsDNA, RNP, Sm, and DNase1L3 were significantly associated with increased activity levels of IFN-I (**figure 3E**). Interestingly, however, antibodies against Ro52Ex4 and Ro52γ — two recently described subsets of anti-Ro52 antibodies^38^ — were linked to increased levels of IFN-I and IFN-II, respectively (**figure 3E**). No autoantibody was associated with increased IFN-III activity (**figure 3E**). Among the individual and combined IFN groups, antibodies to dsDNA, RNP, Sm and, DNaseL3 were predictive of single elevation of IFN-I, as well as co-elevation of the three IFN types (**figure 3F**). Antibodies to Ro52 and Ro52Ex4 were only significantly associated with increased activity levels of IFN-I alone, while antibodies to Ro52γ were the only autoantibodies linked to elevation of IFN-II alone (**figure 3F**). Anti-phospholipid syndrome (APS) antibodies (i.e., anti-β2-glycoprotein I and anti-cardiolipin, anti-B2GPI, and anti-aCL, respectively) were not associated with any IFN type or IFN group. Rather, anti-B2GPI antibodies were protective of single elevated IFN-II (**figure 3F**).

### Transcriptional signatures associated with IFN types in SLE

Patients with SLE display unique blood transcriptional profiles, including a hallmark IFN signature.^2^ To determine whether distinct transcriptional fingerprints are associated with the activity of IFNs in SLE, we used gene expression data from blood samples collected in parallel with the samples used to measure IFN activities. Because clinical subsets in SLE are linked to individual and combined IFN groups (**figure 3D**), we initially performed hierarchical clustering on differentially expressed genes between IFN groups (**figure 1G**) and healthy controls to address whether increased activity of single or combined IFN types is associated with unique transcriptional fingerprints in SLE. Interestingly, this approach failed to identify specific clusters associated with individual or combined IFN groups (**Supplemental figure 2**).

To further define whether specific types of IFN are associated with unique transcriptional profiles in SLE, we performed differential gene expression analysis using a mixed-effects model between SLE samples with high vs. normal IFN for each IFN type. By comparing SLE samples with high vs. normal IFN-I activity, we identified 642 upregulated and 350 downregulated transcripts associated with elevated IFN-I (**figure 4A** and **supplemental file 1**). Enrichment analysis confirmed that the majority of genes related to elevated IFN-I were associated with antiviral response, cell cycle, and immune response (**figure 4B**). Increased activity of IFN-II was associated with 41 upregulated and 51 downregulated transcripts (**figure 4C** and **supplemental file 1**), which are involved in oxidative phosphorylation, NK cell mediated cytotoxicity, mitochondrial gene expression, and RNA metabolism (**figure 4D**). Elevated IFN-III activity was associated with 20 upregulated and 29 downregulated transcripts (**figure 4E** and **supplemental file 1**). Enrichment revealed that most transcripts are involved in hematopoietic stem cell differentiation, complement activation, proteoglycans in cancer, P53 regulation (**figure 4F**).

**Figure. 4.**
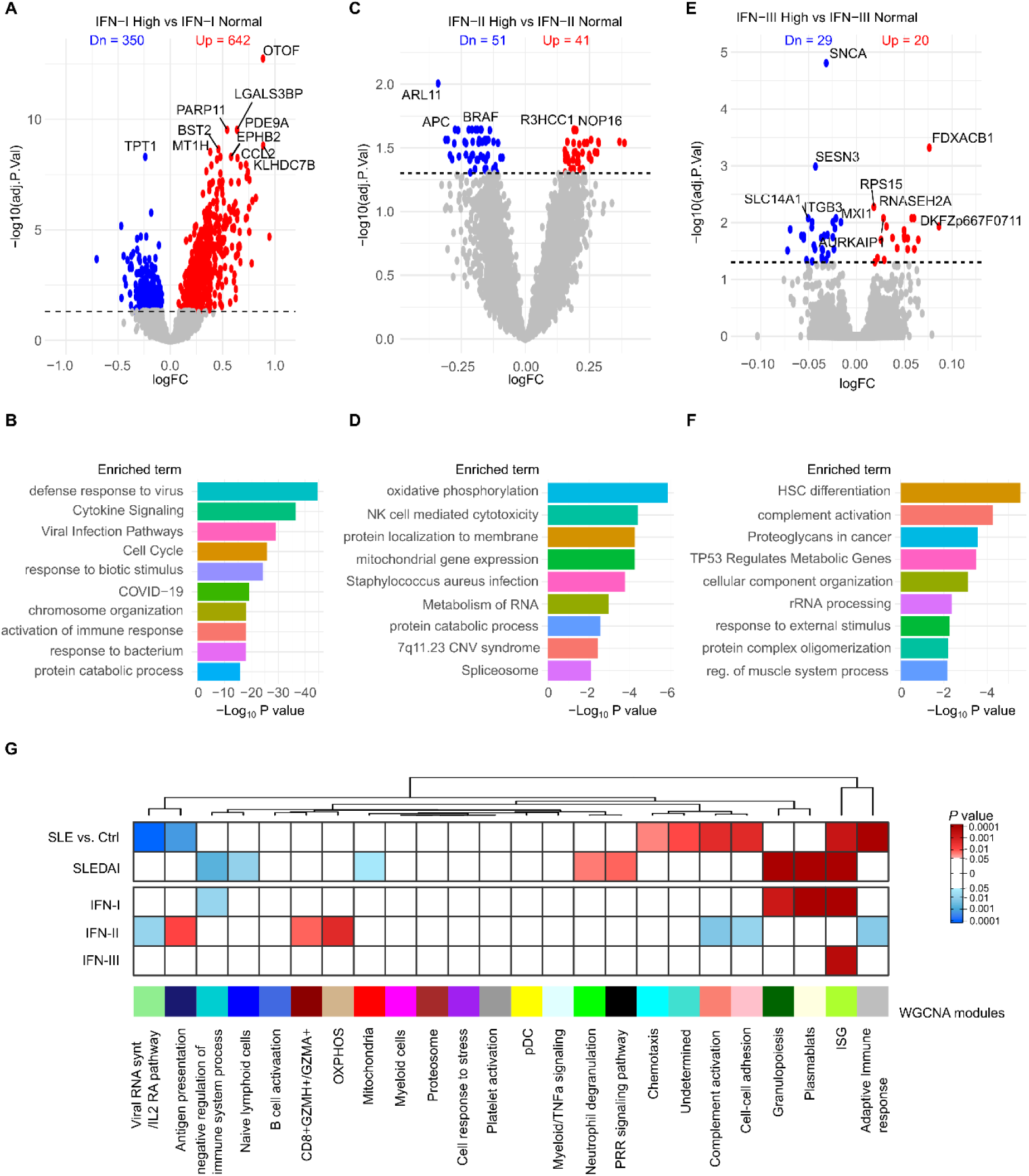
IFN types are associated with distinct transcriptional profiles in SLE. A-F. Differentially expressed transcripts (DET) and enriched terms associated with elevated IFN-I (**A** and **B**), IFN-II (**C** and **D**) and IFN-III (**E** and **F**). Differential expression analysis was done using a linear mixed-effects model to account for patients with repeated samples. Only the top DET were labeled in the volcano plots. The dotted line on each volcano plot equals an adjusted *p* value of 0.05. Enrichment analysis was performed using Metascape.org. Only the top 10 summary terms are shown. (**G**) Association between gene co-expression modules and disease state (SLE vs. Control), disease activity (SLEDA), and activity levels of IFN-I, IFN-II, and IFN-III. Shades of red or blue represent modules positively (β > 0) or negatively associated (β < 0) with the indicated variable, respectively. Associations were done using a linear mixed-effects model.

To gain further insights into the role of IFNs in transcriptional pathways dysregulated in SLE, we performed weighted correlation network analysis (WGCNA)^39^ to define gene co-expression modules using gene expression data from 18 healthy controls and 327 samples from 191 SLE patients. We identified 24 transcriptional modules (**Supplemental file 2**), which were classified according to their association with disease state (i.e., differentially regulated in SLE vs. controls) or disease activity (SLEDAI) (**figure 4G**). Eight modules were associated with disease state (SLE vs. control), of which six were upregulated (adaptive immune response, ISG, cell-cell adhesion, complement, undetermined and chemotaxis), and two were downregulated (antigen presentation and viral RNA synthesis). Seven modules were significantly associated with disease activity, of which five were upregulated (ISG, plasmablasts, granulopoiesis, pattern recognition receptor signaling and neutrophil degranulation), and three downregulated (mitochondria, naïve lymphoid cells and negative regulation of immune system process) (**figure 4G**).

To identify specific modules regulated by IFNs, we modeled the association between IFN types and individual blood transcription modules (**figure 4G**). Interestingly, increased levels of IFN-I, but not IFN-II or IFN-III, were linked with upregulation of disease activity modules (i.e., plasmablasts and granulopoiesis) (**figure 4G**). Instead, IFN-II was associated with upregulation of modules linked to oxidative phosphorylation, CD8^+^*GZMH*^+^ T cells and antigen presentation, and downregulation of adaptive immune response, cell-cell adhesion, complement and viral RNA synthesis modules. IFN-III activity levels were only associated with the ISG module (**figure 4G**).

### Association between the IFN types and the IFN signature

To better understand the relationship between the IFN signature and IFN types in SLE, we performed unsupervised hierarchical clustering using 387 transcripts from the ISG module (**figure 5A**). SLE patients grouped in a gradient of increased expression of ISGs, which paralleled with disease activity and increased levels of IFN-I (**figure 5A**). A common strategy used in clinical trials targeting the IFN-I pathway is to average the expression of four genes (IFI27, IFI44, IFI44L and RSAD2) to classify SLE patients according to IFN activity.^25,40–42^ Using the four-gene IFN signature (4GS), SLE patients are classified as high or low IFN signature when their 4GS was higher than the mean + 2SD compared to healthy controls.^25,40–42^ Using this approach, we confirmed that the 4GS signature strongly correlates with the ISG module (r=0.900, p<0.0001) (**figure 5B**). Paradoxically, however, 4GS showed a poor correlation with IFN-I activity levels (r^2^=0.168, p<0.001) (**figure 5C**). This finding is explained because only 36% (72/201) of SLE samples with a high 4GS have increased activity levels of IFN-I (**figure 5D**). Thus, although it is certain that increased levels of IFN-I are strongly associated with the IFN signature (72/77, 93.5%, p<0.001), up to 64% of samples with high 4GS contain IFN-I activity levels within a normal range. Indeed, the predictive value of 4GS was highly sensitive to detect SLE patients with high IFN-I activity (93%), but its specificity was low (38%). Similarly, 4GS showed low positive and high negative predictive values (35.8% and 96.3%, respectively) to detect increased levels of IFN-I (**figure 5E**). Importantly, increased activity levels of IFN-II and IFN-III were similar in samples with normal or elevated 4GS (IFN-II, p=0.479; IFN-III, p<0.570) (**figure 5D**), implying that neither IFN-II nor IFN-III can explain the expression of the IFN signature in patients with normal activity levels of IFN-I. When we compared clinical and transcriptional features of SLE patients according to 4GS and IFN-I activity (**figure 5F-H**), we found that the subset of patients with increased 4GS and IFN-I activity were characterized by higher SLEDAI, plasmablasts signature and increased activity of IFN-III.

**Figure 5.**
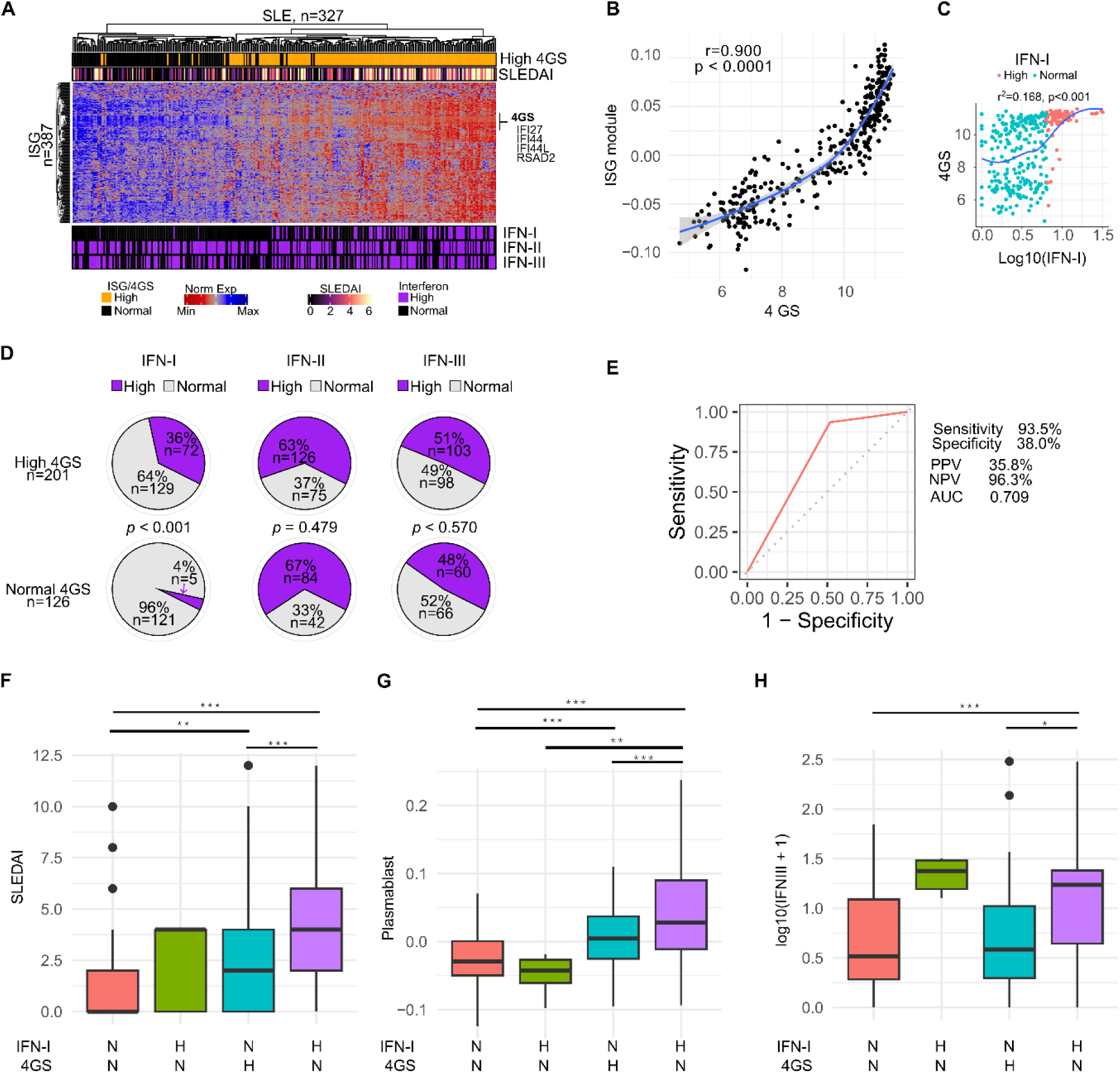
Relationship between IFN-induced gene expression and activity levels of IFNs. **A**. Hierarchical clustering of 387 transcripts from the ISG module defined in Figure 4G. Each column represents an individual patient and each row an individual gene. The top annotations represent the four-gene IFN signature (4GS) as qualitative variable and SLEDAI score. The bottom annotations represent IFN activity levels as dichotomic variables. **B.** Correlation (Person’s r) between the averaged 4GS and the ISG module. **C.** Correlation between the averaged 4GS and log transformed IFN-I activity. **D.** Frequency of normal and increased levels of IFNs according to the 4GS signature. **E.** ROC curve analysis of elevated 4GS signature as classifier of increased levels of IFN-I. **F-H.** Comparison of SLEDAI (**F**), plasmablasts module (**G**), and IFN-III activity (**H**) in SLE patients according to IFN-I and 4GS (N = normal, H = high). Comparisons were done by using a linear mixed-effects linear model in order to account for subjects with repeated samples.

## Discussion

IFNs are promising therapeutic targets in SLE. A deeper understanding of the clinical and pathogenic significance of IFNs in SLE has therefore critical implication for both diagnosis and treatment. To fill gaps in the relationship between IFNs and SLE, we used a novel and convenient assay to precisely measure the independent activity of each IFN type in a large and prospective SLE cohort with extensive clinical, laboratory, and whole blood transcriptional data. Using this approach, we identified novel insights into the potential role of IFNs in SLE pathogenesis and provided a rational explanation for the heterogeneous response in clinical trials targeting IFN-I in SLE. Overall, our data support an essential role of IFNs in SLE pathogenesis and disease activity, but only in specific subsets of patients.

While increased activity levels of IFN-II and IFN-III were twice as common as increased levels of IFN-I in SLE, our data support IFN-I as the master regulator of disease activity in about one-third of patients with SLE. Interestingly, however, disease activity features associated with IFN-I seem to be determined by co-elevation of the other IFN types. Thus, increased activity levels of IFN-I alone were strongly associated with cutaneous lupus, which explains why distinct therapies targeting the IFN-I pathway (either IFN-I producing cells, soluble IFN-I, or the IFN-I receptor) show the most efficacy in SLE patients with cutaneous disease.^40–45^ In contrast, features linked to systemic disease activity – such as arthritis, nephritis and low complement – were associated with increased activity levels of IFN-I plus IFN-II and IFN-III, indicating a synergistic effect of the three IFN families in this severe disease subset.

Interestingly, although co-elevation of IFN-II and IFN-III was associated with a mild increase in SLEDAI, this subgroup is clinically heterogenous, with no association with any particular features of disease activity. Thus, the data point to a model in which co-elevation of IFN-II and IFN-III, the most prevalent combination of IFN types in SLE, creates a low disease activity environment that seems to be amplified by the addition of IFN-I, acting as the determinant to evolve to a more severe disease. In the absence of other IFN types, however, increased activity levels of IFN-I appear to be restricted to cutaneous disease. The finding that hallmark autoantibodies linked to SLE pathogenesis were only associated with disease subsets with high IFN-I activity, as well as the unique association of IFN-I with transcriptional fingerprints linked to disease activity (plasmablasts and granulopoiesis), further support the notion that IFN-I is the main IFN type that dictates the disease outcome in SLE. Nevertheless, the data also anticipate that blocking the IFN-I pathway would be insufficient to treat clinical subsets driven by the synergistic effect of the three IFN types.

An intriguing observation from this study is that, whereas increased activity levels of IFN-I were strikingly associated with the IFN signature, in up to 64% of the cases, the IFN signature was not associated with increased levels of IFN-I. This finding offers a rational explanation for why blocking the IFN-I receptor showed efficacy only in a subset of SLE patients with a high IFN signature,^41^ and highlights that anti-IFN-I therapies are more likely to be effective in patients with high levels of IFN-I rather than a high IFN signature. Similarly intriguing is the finding that neither increased levels of IFN-II nor IFN-III appear to explain the expression of the IFN signature in the subset with normal levels of IFN-I, implying that a different mechanism – likely another cytokine – is responsible for inducing this transcriptional profile. Indeed, not every feature in SLE, such as cytopenias, serositis and APS, was associated with increased levels of IFNs – either individually or collectively –, supporting the notion of IFN-independent subsets in SLE.

While IFN-II alone appears to have a limited role in the IFN signature, as well as disease activity and autoantibodies, and has been shown to be an ineffective therapeutic target in SLE,^46,47^ our data suggest that increased levels of IFN-II are associated with specific transcriptional profiles dysregulated in SLE,^7,8^ including oxidative phosphorylation and the clonal expansion of CD8^+^*GZMH^+^* cells. Moreover, IFN-III seems to contribute to the IFN signature when co-elevated with IFN-I. The interplay of these pathways, together with IFN-I, likely explains the synergistic effects of IFNs in severe SLE.

In summary, our findings underscore that a personalized view of SLE patients according to the activity of specific IFNs may uncover disease subsets amenable for tailored anti-IFN therapies.

## Methods

### Study cohort

We studied 341 plasma/serum samples from 191 SLE patients from the “Study of biological Pathways, Disease Activity and Response markers in patients with Systemic Lupus Erythematosus” (SPARE), and 56 plasma/serum samples from healthy controls. In addition, plasma and serum were collected in parallel from 20 consecutive patients with SLE. SPARE is a prospective observational cohort that has been extensively described previously.^34,35^ Adult patients (age 18 to 75 years-old) who met the definition of SLE per the revised American College of Rheumatology classification criteria^48^ were eligible into the study. At baseline, the patient’s medical history was reviewed, and information on current medications was recorded. The SPARE cohort patients were followed-up over a 2-year period. All patients were treated according to standard clinical practice. Disease activity was assessed using the Safety of Estrogens in Lupus Erythematosus: National Assessment (SELENA) version of the Systemic Lupus Erythematosus Disease Activity Index (SLEDAI)^49^ and physician global assessment (PGA).^50^ C3, C4, anti-dsDNA (Crithidia), complete blood cell count and urinalysis were performed at every visit. All samples were obtained under informed written consent approved by the Johns Hopkins University Institutional Review Board.

### Quantification of activity levels of IFNs

Activity levels of IFN-I, IFN-II and IFN-III were determined in serum/plasma using the HEK-Blue^TM^ IFN-α/β (Cat. # hkb-ifnab), HEK-Blue^TM^ IFN-γ (Cat. # hkb-ifng) and HEK-Blue^TM^ IFN-λ cells (Cat. # hkb-ifnl) from InvivoGen, respectively. Upon activation with IFN, the reporter cells secrete embryonic alkaline phosphatase (SEAP) under the control of the ISG54 promoter. Thus, using a colorimetric substrate, SEAP levels are used to quantify IFN-induced activation in 96-well plates. Briefly, each cell line was plated in 96 well plates and incubated with 100 µl DMEM containing 20% SLE or healthy control heparin/citrate plasma or serum in duplicated. Since recalcification of citrated plasma in DMEM activates the coagulation pathway, 0.6 IU of sodium heparin was added per 100 µl media. To control for heparin, assays performed in serum also contained 0.6 IU of sodium heparin. After 24 hrs at 37°C, 20 μl of supernatant were incubated at room temperature with 180 μl of QUANTI-Blue™ alkaline phosphatase substrate (InvivoGen) for 2 hr to determine the activity of IFN-I and IFN-III, and for 18 hr for IFN-II activity. Using this approach, we found that activity levels of IFNs were similar in serum and plasma collected in parallel (**Supplemental Figure 1**). To quantify the IFN activity, we interpolated the 620 nm absorbance to a standard curve made with serial dilutions of human recombinant IFN-β (PeproTech, Cat. # 300-02BC), IFN-γ (PreproTech, Cat. # 300-02), and IFN-λ3 (IL-28B) (Gibco, Cat. # PHC0894). Cutoff values for IFN-I and IFN-II were determined from healthy control IFN activity using ROC curve analyses setting specificity of 95%. Cutoff value for IFN-III was determined by using a cutoff point above the 95^th^ percentile of IFN-III activity in healthy controls.

### Gene expression analyses

Gene expression analysis from the SPARE cohort was previously described.^35^ CEL files were subjected to RMA background correction, and quantile normalization, using the Oligo package.^2^ To select only expressed genes in whole blood, we filtered out transcripts that had a raw signal < 100 in less than 10% of samples with the genefilter R package. All calculations and analyses were performed using R (ver 4.0.2) and Bioconductor (ver 3.13).^51^ Differentially expressed transcripts (DETs) were analyzed fitting a mixed-effects linear model using dream method from the R package variancePartition.^52^ Functional gene set enrichment analyses were carried out using the online server Metascape.org.^53^ Unsupervised hierarchical clustering with complete linkage was performed by computing a correlation-based distance between genes (Pearson’s method) and the Canberra metric for the distance between subjects. Heatmap visualizations were done using the Complex heatmap R package.^54^ To improve visualization, dendrograms were reordered using the modular leaf ordering methods from the dendsort R package.^55^

### Gene co-expression modules

Gene co-expression modules were determined by weighted correlation network analysis (WGCNA), implemented with the WGCNA R package.^39^ The weighted network was constructed using a thresholding power β of 10 based on the criterion of approximate scale-free topology, a minimum module size of 30, and a medium sensitivity for cluster splitting. Module eigengenes (first principal component) were used as a measure of module activity. Modules were interpreted by gene-set enrichment analysis using Metascape.org.^53^

### Statistical analyses

Comparisons of continuous variables between groups were done using Student’s T test and ANOVA test as indicated. Fisher’s exact test and χ^2^ tests were used for univariate analysis on SPARE cohort variables, as appropriate. The repeatability or intraclass correlation coefficients (ICC), and their corresponding 95% confidence intervals (95% CI), for the IFN types, bodyweight and SLEDAI were estimated using a mixed-effects model implemented with the rptR package.^37^ To estimate the between-patient correlation of disease activity parameters and the IFN types we fitted a Bayesian multivariate multilevel model considering both the IFN type and disease activity as outcomes, and patients with repeated samples as random effects, using the following formula mvbind (y, x) ∼ 1 + (1|c|patient) using the brms package.^56^ Associations of clinical variables with continuous levels of the IFN types were determined by mixed-effects models to consider group asymmetry and the effect of subjects with longitudinal samples using the lme4 package.^57^ Associations of variables with IFN type subsets as dependent variables were determined by fitting multinomial log-linear models via neural networks considering patients with repeated samples as a random effect using the R package nnet.^58^ Statistical significance was set at p < 0.05. The statistical analyses were carried with the R software version 4.0.2.

## Acknowledgements

Funding for this project was provided by the Jerome L. Greene Foundation, the National Institute of Allergy and Infectious Diseases (NIAID) and the National Institute of Arthritis and Musculoskeletal and Skin Diseases (NIAMS) at the National Institutes of Health (NIH) grants number R21 AI147598, R21 AI169851, R21 AI176766 and R01 AR069572. The content of this paper is solely the responsibility of the author and does not represent the official views of the NIAMS, NIAID or the NIH.

## Author Contributions

Conception and design: E.G.B. and F.A. Designed experiments: E.G.B. and F.A. Performed experiments and statistical analyses: E.G.B. and VA. Data acquisition and analyses: E.G.B., D.G., V.A., E.D., M.P., and FA. Interpretation of data: E.G.B., E.D., M.P., and F.A. Data curation and processing: E.G.B. Writing—original draft: E.G.B. and F.A. Writing—review and editing: E.G.B., D.G., V.A., E.D., M.P., and FA. All authors reviewed, edited, and approved the manuscript.

## Data availability

Microarray data are available from the Gene Expression Omnibus under accession number GSE45291 and GSE121239.

