## Supplementary Materials for "Uncoupling interferons and the interferon signature explain clinical and transcriptional subsets in SLE"

**Contents**

**Supplemental figure 1.** Correlation between activity levels of IFN-I, IFN-II and IFN-III in serum and plasma using HEK-Blue$^{TM}$ cells.

**Supplemental Figure 2.** Individual or combined IFN type groups are not associated with specific transcriptomes in SLE.

**Supplemental table 1.** Demographic characteristics of SLE patients from SPARE

**Supplemental table 2.** Association between IFN-I activity, disease duration and disease activity.

**Supplemental table 3.** Association between IFN-II activity, disease duration and disease activity.

**Supplemental table 4.** Association between IFN-III activity, disease duration and disease activity.

**Supplemental table 5.** Associations between clinical traits captured at time of visit and activity levels of IFN types.

**Supplemental table 6.** Associations between SLE disease activity features and individual and combined IFN groups.

**Supplemental file 1 (Excel file)**

**Supplemental file 2 (Excel file)**

### Supplemental Figures and Tables

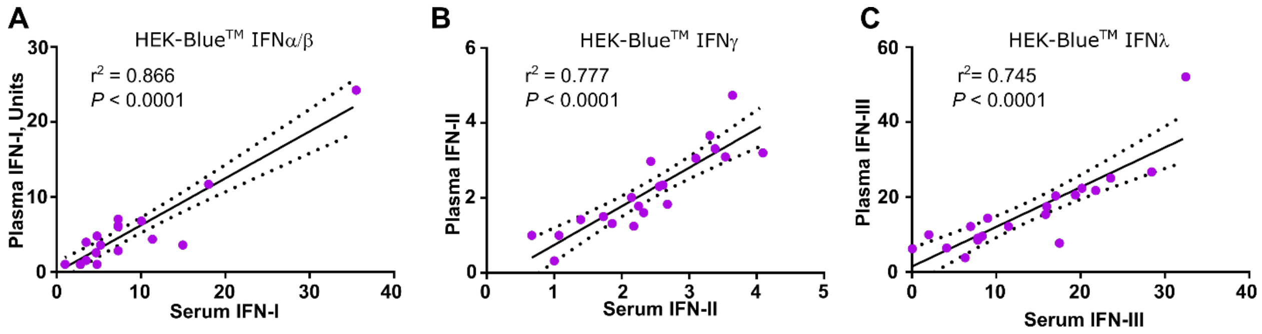


**Supplemental figure 1.** Correlation between activity levels of IFN-I, IFN-II and IFN-III in serum and plasma using HEK-Blue™ cells. (**A-C**) HEK-Blue™ IFN-α/β (**A**), HEK-Blue™ IFN-γ (**B**) and HEK-Blue™ IFN-λ (**C**) were incubated with serum or plasma collected in parallel from 20 consecutive patients with SLE. $r^2$ was calculated using a linear regression model with plasma IFN activity as dependent variable.

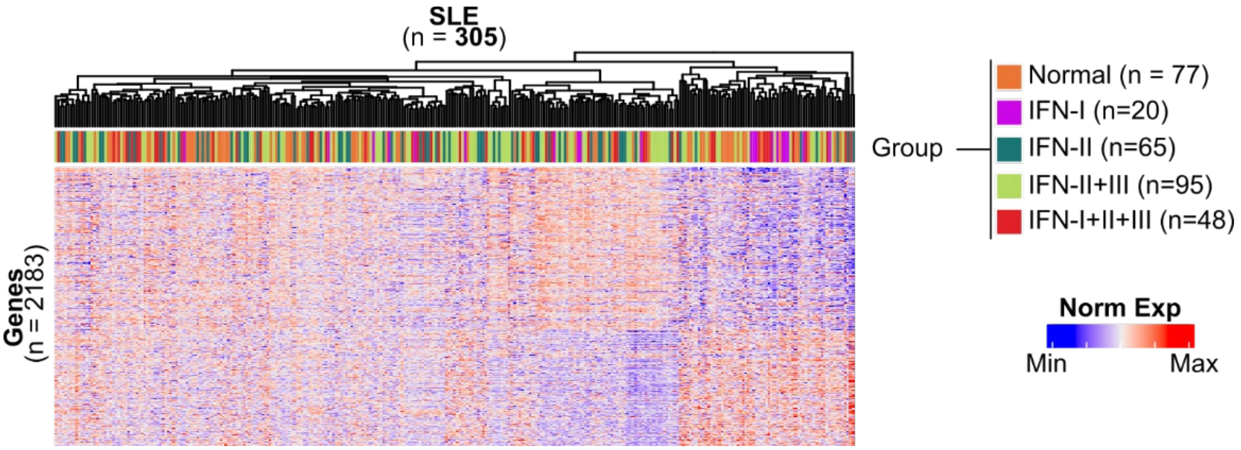


**Supplemental figure 2.** Individual or combined IFN type groups are not associated with specific transcriptomes in SLE. Unsupervised hierarchical clustering of 2183 differentially expressed transcripts between healthy controls and SLE patients according to individual or combined groups of IFN types (n=305). Each column represents an individual patient and each row an individual gene. The top annotations represent the IFN group.

**Supplemental Table 1.** Demographic characteristics of SLE patients from SPARE

| **Variable** | **n** | **n = 190** |
|---|---|---|
| Female Sex | 190 | 177 (93%) |
| Race | 190 | |
| White | | 100 (53%) |
| Black | | 74 (39%) |
| Asian | | 9 (4.7%) |
| Other | | 7 (3.7%) |
| Smoking | 190 | 15 (7.9%) |
| SLEDAI | 187 | 2 (0, 15)[1] |
| Renal SLE | 190 | 99 (52%) |
| Sjogren's Syndrome | 190 | 48 (25%) |
| Anti-DNA | 190 | 123 (65%) |
| Anti-Sm | 189 | 38 (20%) |
| Anti-Ro52 | 190 | 75 (39%) |
| Anti-La | 189 | 28 (15%) |
| Anti-RNP | 189 | 52 (28%) |
| Anti-Ro52$_{EX4}$ | 190 | 95 (50%) |
| Anti-Ro52γ | 190 | 41 (22%) |
| Anti-DNase1L3 | 156 | 47 (30%) |
| Current treatment | | |
| Prednisone | 190 | 68 (36%) |
| Hydroxicholoroquine | 190 | 167 (88%) |
| Cytotoxic treatment | 190 | 117 (62%) |

[1]Median (min-max). Cytotoxic treatment includes Cyclophosphamide, Mycophenolic acid, Azathioprine, and Methotrexate.

**Supplemental table 2.** Association between IFN-I activity, disease duration and disease activity.

| | IFN-I, AU/mL | | |
|---|---|---|---|
| | Model 1<br>β (95% CI) | Model 2<br>β (95% CI) | Model 3<br>β (95% CI) |
| Disease duration, yrs | -0.004 (-0.007, -0.001) | | -0.004 (-0.007, -0.001) |
| | p = 0.020 | | p = 0.021 |
| SLEDAI | | 0.027 (0.016, 0.037) | 0.026 (0.016, 0.036) |
| | | p <0.0001 | p <0.0001 |
| Constant | 0.630 (0.555, 0.705) | 0.485 (0.440, 0.531) | 0.557 (0.481, 0.632) |
| | p <0.0001 | p <0.0001 | p <0.0001 |
| Observations | 322 | 322 | 322 |

Associations were determined using a mixed-effects linear model.

**Supplemental table 3**. Association between IFN-II activity, disease duration and disease activity.

| | IFN-II, AU/mL | | |
|---|---|---|---|
| | Model 1<br>β (95% CI) | Model 2<br>β (95% CI) | Model 3<br>β (95% CI) |
| Disease duration, yrs | 0.003 (-0.00001, 0.005) | | 0.003 (0.0001, 0.005) |
| | p = 0.051 | | p = 0.041 |
| SLEDAI | | 0.006 (-0.003, 0.015) | 0.007 (-0.002, 0.016) |
| | | p = 0.172 | p = 0.134 |
| Constant | 0.643 (0.583, 0.703) | 0.678 (0.639, 0.717) | 0.623 (0.558, 0.688) |
| | p <0.0001 | p <0.0001 | p <0.0001 |
| Observations | 322 | 322 | 322 |

Associations were determined using a mixed-effects linear model.

**Supplemental table 4.** Association between IFN-III activity, disease duration and disease activity.

| | IFN-III, AU/mL | | |
|---|---|---|---|
| | Model 1<br>β (95% CI) | Model 2<br>β (95% CI) | Model 3<br>β (95% CI) |
| Disease duration, yrs | 0.003 (-0.002, 0.008) | | 0.004 (-0.002, 0.009) |
| | p = 0.246 | | p = 0.169 |
| SLEDAI | | 0.027 (0.008, 0.046) | 0.028 (0.009, 0.047) |
| | | p = 0.005 | p = 0.004 |
| Constant | 0.698 (0.581, 0.816) | 0.688 (0.609, 0.767) | 0.616 (0.488, 0.745) |
| | p <0.0001 | p <0.0001 | p <0.0001 |
| Observations | 322 | 322 | 322 |

Associations were determined using a mixed-effects linear model.

**Supplemental table 5.** Associations between clinical traits captured at time of visit and activity levels of IFN types.

| | IFN types (dependent variables) | | | | | |
|---|---|---|---|---|---|---|
| | IFN-I | | IFN-II | | IFN-III | |
| Variable (Independent) | β | *p value* | β | *p value* | β | *p value* |
| Systolic BP | -0.007(-0.033,0.018) | 0.567 | 0.004(-0.019,0.026) | 0.753 | -0.014(-0.195,0.167) | 0.881 |
| Diastolic BP | 0.006(-0.037,0.049) | 0.779 | 0.038(0.001,0.075) | **0.044** | -0.015(-0.318,0.285) | 0.920 |
| Race | 1.046(-1.715,3.803) | 0.677 | 1.452(-0.81,3.712) | 0.281 | 4.58(-12.314,21.421) | 0.660 |
| Sex | 0.094(-2.184,2.376) | 0.935 | -0.025(-1.948,1.901) | 0.980 | -0.119(-15.373,15.147) | 0.988 |
| LAI score | 0.914(0.312,1.521) | **0.003** | 0.071(-0.459,0.601) | 0.791 | 5.058(0.714,9.401) | **0.023** |
| A: Fatigue | -0.047(-0.093,-0.001) | NA | 0.032(-0.006,0.07) | NA | 0.008(-0.278,0.294) | NA |
| B: Rash | 1.429(0.505,2.358) | **0.003** | -0.48(-1.295,0.335) | 0.247 | -2.039(-8.79,4.698) | 0.552 |
| C: Joints | -0.688(-1.701,0.323) | 0.182 | 0.307(-0.602,1.214) | 0.507 | 2.698(-4.89,10.286) | 0.485 |
| D: Serositis | -0.3(-3.048,2.422) | 0.829 | -1.077(-3.569,1.409) | 0.395 | -1.162(-22.776,20.423) | 0.916 |
| A: Neurological | -0.629(-3.271,2.013) | 0.639 | -0.395(-2.629,1.839) | 0.728 | 4.451(-13.312,22.221) | 0.622 |
| B: Renal | 0.795(0.025,1.561) | **0.043** | 0.614(-0.046,1.278) | 0.068 | 7.177(2.065,12.301) | **0.006** |
| C: Pulmonary | NA | NA | NA | NA | NA | NA |
| D: Hematological | 1.716(-0.016,3.449) | 0.052 | -1.202(-2.704,0.304) | 0.117 | -1.166(-13.424,11.096) | 0.852 |
| SLEDAI | 0.295(0.154,0.437) | **< 0.001** | 0.057(-0.066,0.18) | 0.363 | 0.837(-0.188,1.86) | 0.109 |
| *Neurological** | | | | | | |
| Organic brain syndrome | -0.674(-6.321,4.977) | 0.815 | -0.343(-5.119,4.436) | 0.888 | 0.022(-37.992,38.044) | 0.999 |
| Cranial nerve disorder | 0.207(-7.764,8.179) | 0.959 | 0.012(-6.728,6.755) | 0.997 | 29.593(-23.966,83.161) | 0.278 |
| Vasculitis | -0.719(-3.079,1.64) | 0.549 | 0.124(-2.006,2.267) | 0.990 | -16.275(-34.258,1.682) | 0.076 |
| *Renal* | | | | | | |
| Urinary casts | NA | NA | NA | NA | NA | NA |
| Hematuria | 0.895(-1.376,3.168) | 0.439 | 1.469(-0.546,3.481) | 0.152 | -3.018(-19.642,13.613) | 0.721 |
| Proteinuria | 1.826(0.254,3.408) | **0.023** | 0.725(-0.69,2.144) | 0.314 | 18.118(6.281,29.958) | **0.003** |
| Pyuria | -0.546(-2.699,1.616) | 0.619 | 0.952(-0.969,2.875) | 0.331 | -0.368(-16.601,15.864) | 0.965 |
| Arthritis | 0.584(-1.009,2.183) | 0.472 | 1.801(0.385,3.217) | **0.013** | 3.051(-8.932,15.034) | 0.617 |
| Myositis | NA | NA | NA | NA | NA | NA |
| *Immunological* | | | | | | |
| Low Complement | 3.069(1.947,4.189) | **< 0.001** | -0.489(-1.502,0.521) | 0.341 | 10.003(2.028,17.953) | **0.014** |
| Increased DNA binding | 2.982(1.917,4.043) | **0.001** | -0.656(-1.605,0.291) | 0.174 | 5.9(-1.361,13.196) | 0.111 |
| *Cutaneous* | | | | | | |
| Rash | 2.836(1.138,4.534) | **0.001** | 0.006(-1.502,1.517) | 0.994 | 2.121(-10.02,14.274) | 0.731 |
| Alopecia | 0.709(-0.367,1.783) | 0.196 | 0.088(-0.843,1.019) | 0.853 | 1.848(-5.707,9.347) | 0.630 |
| Mucosal ulcers | -3.415(-5.94,-0.875) | **0.009** | -1.972(-4.226,0.287) | 0.087 | -9.478(-28.385,9.431) | 0.325 |
| *Serositis* | | | | | | |
| Pleurisy | -0.723(-4.084,2.611) | 0.671 | -1.346(-4.388,1.694) | 0.384 | -5.799(-32.118,20.501) | 0.665 |
| Pericarditis | NA | NA | NA | NA | NA | NA |
| *Hematological* | | | | | | |
| Thrombocytopenia | 0.537(-4.085,5.162) | 0.819 | -2.424(-6.322,1.475) | 0.222 | -5.797(-36.888,25.303) | 0.714 |
| Leukopenia | 2.97(-0.089,6.029) | 0.057 | -1.76(-4.446,0.927) | 0.198 | 2.973(-19.033,24.98) | 0.791 |
| *Infections* | | | | | | |
| Viral infection | -1.475(-3.595,0.644) | 0.179 | 0.765(-1.145,2.669) | 0.431 | -5.038(-21.183,11.102) | 0.540 |
| Bacterial infection | -0.347(-1.814,1.123) | 0.643 | 1.371(0.072,2.67) | **0.039** | 4.773(-6.131,15.676) | 0.390 |
| Thrush | 1.275(-1.897,4.519) | 0.433 | 1.517(-1.267,4.293) | 0.284 | 54.195(31.002,77.448) | **< 0.001** |
| *Treatment* | | | | | | |
| Cytotoxic | 0.528(-0.514,1.557) | 0.318 | -0.302(-1.164,0.558) | 0.489 | 6.144(-0.386,12.626) | 0.065 |
| Hydroxychloroquine | 0.127(-1.106,1.36) | 0.839 | -0.545(-1.586,0.497) | 0.304 | -6.154(-14.406,1.99) | 0.139 |
| NSAID use | -1.222(-2.426,-0.011) | **0.048** | -0.249(-1.268,0.774) | 0.631 | 0.376(-7.411,8.24) | 0.925 |
| Clopidogrel | 0.082(-2.141,2.312) | 0.942 | 0.034(-1.849,1.922) | 0.972 | -6.901(-21.626,7.841) | 0.358 |
| Antihypertensive drugs | -0.421(-1.378,0.535) | 0.387 | 0.476(-0.351,1.304) | 0.258 | 1.955(-4.667,8.587) | 0.562 |
| Diuretic | -1.171(-2.212,-0.129) | **0.028** | 0.005(-0.885,0.894) | 0.991 | -2.61(-9.458,4.251) | 0.454 |
| Calcium antagonists | 0.141(-1.152,1.43) | 0.830 | 0.015(-1.074,1.104) | 0.978 | 7.232(-1.093,15.594) | 0.088 |
| Statin | -0.195(-1.266,0.877) | 0.721 | 0.234(-0.676,1.141) | 0.613 | -1.923(-8.898,5.046) | 0.587 |

β and p values were calculated using a mixed-effect model to control for the effect of patients with repeated samples and adjusted by disease duration. BP: blood pressure. LAI: Lupus activity index. *Other neurological items evaluated in the SELENA-SLEDAI such as seizures, psychosis, visual disturbance, lupus headache, and cerebrovascular accidents where not present in any patient at time of visit. Cytotoxic drugs were defined as: Leflunomide, Mycophenolate, Chlorambucil, Cyclosporin, Cytoxan, Tacrolimus, Azathioprine, Methotrexate, Rituximab, Etanercept, Abatacept or Adalimumab.

**Supplemental table 6.** Associations between SLE disease activity features and individual and combined IFN groups.

| Predictor | IFN group | OR (95% CI) | P value |
|---|---|---|---|
| SLEDAI | **IFN-I** | **1.38 (1.16,1.63)** | **< 0.001** |
| | IFN-II | 1.08 (0.94,1.25) | 0.291 |
| | **IFN-II + III** | **1.17 (1.03,1.33)** | **0.016** |
| | **IFN-I + II + III** | **1.38 (1.2,1.58)** | **< 0.001** |
| Vasculitis | IFN-I | 3.89 (0.23,65.17) | 0.344 |
| | IFN-II | 5.02 (0.55,46.1) | 0.154 |
| | IFN-II + III | 3.29 (0.36,30.07) | 0.292 |
| | **IFN-I + II + III** | **0 (0,0)** | **< 0.001** |
| Hematuria | IFN-I | 1.92 (0.17,22.32) | 0.602 |
| | IFN-II | 1.2 (0.16,8.74) | 0.860 |
| | IFN-II + III | 1.2 (0.2,7.39) | 0.842 |
| | IFN-I + II + III | 1.59 (0.22,11.65) | 0.650 |
| Any Renal | IFN-I | 1.56 (0.28,8.68) | 0.615 |
| | IFN-II | 0.7 (0.16,3.05) | 0.635 |
| | IFN-II + III | 2.45 (0.84,7.15) | 0.101 |
| | **IFN-I + II + III** | **3.23 (1.01,10.32)** | **0.048** |
| Proteinuria | IFN-I | 1.92 (0.17,22.33) | 0.602 |
| | IFN-II | 0.59 (0.05,6.65) | 0.668 |
| | IFN-II + III | 3.4 (0.7,16.49) | 0.130 |
| | **IFN-I + II + III** | **5.21 (1.01,27.01)** | **0.049** |
| Pyuria | **IFN-I** | **0 (0,0)** | **< 0.001** |
| | IFN-II | 0.59 (0.05,6.65) | 0.668 |
| | IFN-II + III | 1.62 (0.29,9.11) | 0.583 |
| | IFN-I + II + III | 2.43 (0.39,15.13) | 0.340 |
| Arthritis | IFN-I | 1.92 (0.17,22.32) | 0.602 |
| | IFN-II | 2.47 (0.44,13.97) | 0.305 |
| | IFN-II + III | 2.94 (0.59,14.57) | 0.188 |
| | **IFN-I + II + III** | **6.23 (1.24,31.39)** | **0.027** |
| Low Complement | IFN-I | 2.44 (0.72,8.35) | 0.154 |
| | IFN-II | 0.63 (0.2,1.99) | 0.434 |
| | IFN-II + III | 0.97 (0.38,2.48) | 0.953 |
| | **IFN-I + II + III** | **6.75 (2.75,16.55)** | **< 0.001** |
| Increased DNA binding | **IFN-I** | **12 (3.76,38.27)** | **< 0.001** |
| | IFN-II | 0.75 (0.31,1.82) | 0.531 |
| | IFN-II + III | 0.76 (0.34,1.67) | 0.495 |
| | **IFN-I + II + III** | **5.14 (2.3,11.48)** | **< 0.001** |
| Rash | **IFN-I** | **18.5 (1.94,176.77)** | **0.011** |
| | IFN-II | 5.02 (0.55,46.1) | 0.154 |
| | IFN-II + III | 3.29 (0.36,30.07) | 0.292 |
| | IFN-I + II + III | 6.73 (0.73,62.13) | 0.093 |
| Alopecia | **IFN-I** | **3.56 (1.24,10.24)** | **0.018** |
| | IFN-II | 1.13 (0.49,2.63) | 0.772 |
| | IFN-II + III | 1.33 (0.63,2.82) | 0.456 |
| | IFN-I + II + III | 1.62 (0.68,3.83) | 0.274 |

| Predictor | IFN group | OR (95% CI) | P value |
|---|---|---|---|
| Mucosal Ulcers | **IFN-I** | **0 (0,0)** | **< 0.001** |
| | IFN-II | 1.2 (0.23,6.17) | 0.827 |
| | IFN-II + III | 0.26 (0.03,2.53) | 0.245 |
| | **IFN-I + II + III** | **0 (0,0)** | **< 0.001** |
| Pleurisy | **IFN-I** | **0 (0,0)** | **< 0.001** |
| | IFN-II | 1.19 (0.07,19.48) | 0.901 |
| | IFN-II + III | 0.8 (0.05,12.94) | 0.872 |
| | **IFN-I + II + III** | **0 (0,0)** | **< 0.001** |
| Thrombocytopenia | IFN-I | 3.9 (0.23,65.18) | 0.344 |
| | **IFN-II** | **0 (0,0)** | **< 0.001** |
| | IFN-II + III | 0.8 (0.05,12.94) | 0.872 |
| | **IFN-I + II + III** | **0 (0,0)** | **< 0.001** |
| Leukopenia | **IFN-I** | **0 (0,0)** | **< 0.001** |
| | **IFN-II** | **0 (0,0)** | **< 0.001** |
| | **IFN-II + III** | **0 (0,0)** | **< 0.001** |
| | IFN-I + II + III | 1.04 (0.17,6.49) | 0.964 |
| Cytotoxic Use | **IFN-I** | **4.03 (1.33,12.24)** | **0.014** |
| | IFN-II | 1.01 (0.51,1.98) | 0.982 |
| | IFN-II + III | 0.91 (0.49,1.69) | 0.769 |
| | **IFN-I + II + III** | **2.24 (1.07,4.7)** | **0.033** |
| Plaquenil | IFN-I | 0.41 (0.11,1.58) | 0.196 |
| | IFN-II | 0.55 (0.19,1.53) | 0.249 |
| | **IFN-II + III** | **0.27 (0.11,0.66)** | **0.004** |
| | **IFN-I + II + III** | **0.35 (0.12,0.97)** | **0.043** |

Associations were determined by a multinomial log-linear model via neural networks considering patients with repeated samples as a random effect.
